# Plasma proteome analysis of patients with hereditary transthyretin-mediated (hATTR) amyloidosis establishes neurofilament light chain (NfL) as a biomarker of disease and treatment response

**DOI:** 10.1101/19011155

**Authors:** Simina Ticau, Gautham V Sridharan, Shira Tsour, William L Cantley, Amy Chan, Jason A Gilbert, David Erbe, Emre Aldinc, Mary M Reilly, David Adams, Michael Polydefkis, Kevin Fitzgerald, Akshay Vaishnaw, Paul Nioi

**Author notes:** Correspondence to: Paul Nioi, Alnylam Pharmaceuticals, 675 W Kendall St, Cambridge, MA 02142, USA. These authors contributed equally to this work (Simina Ticau, Gautham V Sridharan).

## Abstract

**Background:** Hereditary transthyretin-mediated (hATTR) amyloidosis is a rare, progressively debilitating, and fatal disease caused by deposition of aggregated transthyretin amyloid in multiple organs and tissues. Highly variable disease penetrance has made it difficult to predict disease onset and progression. Clinically validated, non-invasive plasma biomarkers may facilitate earlier diagnosis and aid monitoring of disease progression.

**Methods:** Plasma levels of >1000 proteins were measured in patients with hATTR amyloidosis with polyneuropathy who received either placebo or patisiran in the phase 3 APOLLO study (NCT01960348) and in a cohort of healthy individuals. The impact of patisiran treatment on the time profile of each protein was determined by a linear mixed model at 0, 9, and 18 months. Neurofilament light chain (NfL) protein was further assessed using an orthogonal quantitative approach.

**Findings:** A significant change in the levels of 66 proteins was observed with patisiran *vs* placebo, with change in NfL, a marker of neuronal damage, most significant (p<10^−20^). Analysis of the changes in protein levels demonstrated that the proteome of patients treated with patisiran trended towards healthy individuals at 18 months. Plasma NfL levels in healthy controls were four-fold lower than in patients with hATTR amyloidosis with polyneuropathy (16·3 [SD 12·0] pg/mL *vs* 69·4 [SD 42·1] pg/mL, p<10^−16^). Levels of NfL at 18 months increased with placebo (99·5 [SD 60·1] pg/mL) and decreased with patisiran treatment (48·8 [SD 29·9] pg/mL). At 18 months, improvement in modified Neuropathy Impairment Score+7 (mNIS+7) in patisiran-treated patients significantly correlated with a reduction in NfL levels (*R*=0·43, p<10^−7^).

**Interpretation:** The observed correlation of NfL reduction with patisiran treatment and improvement in mNIS+7 suggests it may serve as a biomarker of nerve damage and polyneuropathy in hATTR amyloidosis. This biomarker may enable earlier diagnosis of polyneuropathy in patients with hATTR amyloidosis and facilitate monitoring of disease progression.

## Introduction

Hereditary transthyretin-mediated (hATTR) amyloidosis is a rare, progressively debilitating, and fatal disease caused by pathogenic mutations in the transthyretin (*TTR*) gene that result in accumulation of amyloid fibrils throughout the body including in the heart, peripheral nerves, and gastrointestinal tract.^1,2^ This accumulation leads to damage of organs and tissue that can include peripheral sensorimotor neuropathy, autonomic neuropathy, and cardiomyopathy, leading to decreased quality of life and eventually death.^3^ The majority of patients with hATTR amyloidosis develop a mixed phenotype of both polyneuropathy and cardiomyopathy as a result of this multisystem involvement.^4-6^ hATTR amyloidosis affects approximately 50 000 people worldwide and has a median survival of 4·7 years following diagnosis,^7^ with a reduced survival of 3·4 years for patients presenting with cardiomyopathy.^8^ Due to variability of the initially affected tissues, age of onset, and penetrance of hATTR amyloidosis, it is difficult to predict disease onset and progression in individual patients. Thus, there is a need for improved understanding of the disease as a whole, including the ability to identify, monitor, and effectively treat patients.

Diagnosis of hATTR amyloidosis remains challenging, including identification of patients with manifest disease, and ascertaining if and when asymptomatic carriers with pathogenic *TTR* mutations become symptomatic.^9,10^ Penetrance of the disease in carriers varies widely by region^11^ and *TTR* mutation.^7^ Additionally, since TTR fibrils aggregate and deposit in tissues over time, subclinical nerve damage is likely to occur prior to presentation of overt symptomatology. One measure used to assess severity and progression of polyneuropathy in patients with hATTR amyloidosis is the modified Neuropathy Impairment Score +7 (mNIS+7).^12^ mNIS+7 is a composite score (range: 0–304) which assesses sensory, motor, and autonomic nervous system functions, with a higher mNIS+7 score indicating worse neuropathy. While mNIS+7 is considered a helpful tool, it can be burdensome to administer, so advancements are necessary to improve our ability to diagnose hATTR amyloidosis earlier and assess its severity with greater objectivity. Discovery of novel plasma biomarkers associated with disease progression could potentially provide minimally invasive measures that facilitate earlier patient diagnosis, improved monitoring of disease progression, and response to therapeutic intervention.

Due to the low prevalence of hATTR amyloidosis, there has been a limited number of clinical proteomic studies conducted in hATTR amyloidosis patient samples. A study was conducted to investigate plasma protein profiles in patients presenting with hATTR amyloidosis relative to healthy controls; however, the sample size was relatively small.^13,14^ The pivotal phase 3 APOLLO study (NCT01960348) was a randomised, double-blind, placebo-controlled trial of patisiran, a lipid nanoparticle-delivered RNA interference (RNAi) therapeutic that reduces serum TTR levels by inhibiting hepatic synthesis of the disease-causing mutant and wild-type (wt) TTR proteins.^6,15^ In APOLLO, patisiran-treated patients experienced improvements in the primary endpoint (mNIS+7) and all secondary endpoints with an acceptable benefit:risk profile.^6^ The objective of this study was to use plasma samples collected during APOLLO to evaluate the change in circulating proteins in response to patisiran treatment in patients with hATTR amyloidosis with polyneuropathy, with the aim of identifying potential clinical biomarkers of hATTR amyloidosis that correlate with treatment response. The clinical utility of biomarkers could conceivably include monitoring of disease progression and treatment effect; biomarker development could potentially lead to an early diagnostic parameter or predictor of when disease may become manifest in asymptomatic carriers of pathogenic *TTR* mutations.

## Methods

### Study design and participants

The present study includes analysis of plasma samples in a subset of patients enrolled in the APOLLO study. During the APOLLO study patients received either patisiran 0·3 mg/kg or placebo intravenously every 3 weeks for 18 months.^6^ Patients were included in this analysis if they completed the APOLLO study and had plasma samples available at baseline and 9 and 18 months for biomarker assessment. Healthy control samples (n=57) were collected under IRB-approved protocols separately and were age-, sex-, and race-matched to the baseline demographics of the APOLLO patients included in this analysis (Dx Biosamples, LLC, San Diego, CA).

### Plasma measurement of biomarkers

Plasma samples of patients enrolled in APOLLO were collected according to the study protocol prior to dosing.^6^ Levels of 1161 proteins were measured using 13 human panels containing 92-plex immunoassays (Olink Proteomics, Watertown, MA) with intra-assay coefficients of variation ranging from 2% to 14% and inter-assay coefficients of variation ranging from 5% to 26%. Olink^®^ uses a proprietary proximity extension assay technology to combine a detection step involving oligonucleotide-labelled antibodies with a proximity-dependent DNA polymerisation step and a real-time quantitative PCR amplification step to measure relative levels of multiple biomarkers simultaneously. Relative protein levels are reported as normalised protein expression (NPX) values which are on a log_2_ scale.

Subsequent quantitative measurements of neurofilament light chain (NfL) were made using an ultrasensitive single molecule array method, in duplicate,^16^ in a subset of patients where sufficient volume of plasma was available (n=112 patisiran-treated, n=47 placebo-treated), at baseline and 18 months. All NfL values were within the linear ranges of the assay except for two measurements, which were excluded.

### Statistical analysis

To determine which protein levels changed the most over time as a result of placebo *vs* patisiran treatment, a linear mixed model regression analysis was performed for each protein to determine if there was a significant differential time profile based on treatment. The Wilkinson notation representation of the model is given by: “Protein NPX∼Treatment+Time+Treatment:Time+Age+Sex+(1|Patient)”, where the response Protein NPX represents protein levels, Treatment denotes whether the patient was administered placebo or patisiran, Time is a numeric quantity representing 0, 9, or 18 months, Treatment:Time is the interaction term, age/sex are additional covariates, and individual patients (Patient) are assigned random intercepts. The model was regressed for each protein using the fit linear mixed-effects model (fitlme) function (Matlab 9.3) and the coefficients and p values associated with the Treatment:Time interaction coefficient were stored and plotted.

Principal component analysis is a statistical procedure used to convert a set of observations that may be correlated into a set of linearly uncorrelated values, called principal components. This transformation results in the first principal component accounting for the largest possible variance in the data, with each subsequent principal component accounting for less variance. Here the analysis is used to display the levels of 66 proteins in various cohorts of individuals (healthy control, baseline APOLLO, APOLLO/placebo, and APOLLO/patisiran) on a simple two-dimensional plot by plotting the two principal components that explain most of the variability in the data.

All box plots are based on standard metrics, namely the minimum value, maximum value, lower and upper quartiles, and median value. Outliers are denoted as those values that are more than 1·5 times the interquartile range) above the third quartile or below the first quartile. Plots were made using the Python Matplotlib graphics environment. All correlation analysis was performed using Pearson’s correlation coefficient and all comparisons of means between the two groups were performed using the student t-test in R. Principal component analysis was performed using Python’s sklearn package, and the 99·9% confidence intervals were drawn using the variance–covariance matrix of each treatment group’s multivariate distribution using the Ellipse patch as part of Matplotlib.

## Results

In order to compare proteomes, subjects comprised 136 patisiran-treated and 53 placebo-treated patients with hATTR amyloidosis with polyneuropathy from the APOLLO study, and 57 healthy controls that were age-, sex-, and race-matched to the APOLLO patients (figure 1; table 1). The APOLLO patient cohort was demographically representative of the patients enrolled in APOLLO.^6^

**Table 1:** Baseline demographics of APOLLO study patients participating in the biomarker study and the age-/gender-/race-matched healthy controls

|  | APOLLO |  |  | Healthy controls |  |
| --- | --- | --- | --- | --- | --- |
|  | Placebo (baseline) | Patisiran (baseline) | Total |  | p |
|  | n=53 | n=136 | n=189 | n=57 | value |
| Mean age in years (SD) | 62.4 (11.0) | 59.7 (12.1) | 60.5 (11.8) | 58.6 (12.1) | 0.32 |
| Sex (% male) | 41 (77.4) | 101 (74.3) | 142 (75.1) | 42 (73.7) | 0.93 |
| Race (%) |  |  |  |  |  |
| Asian | 15 (28.3) | 23 (16.9) | 38 (20.1) | 12 (21.1) | 0.90 |
| Black or African American | 0 (0) | 4 (2.9) | 4 (2.1) | 2 (3.5) | 0.56 |
| White | 37 (69.8) | 105 (77.2) | 142 (75.1) | 43 (75.4) | 0.99 |
| Multiple/other/unknown | 1 (1.9) | 4 (2.9) | 5 (2.6) | 0 (0) | 0.22 |

**Figure 1:**
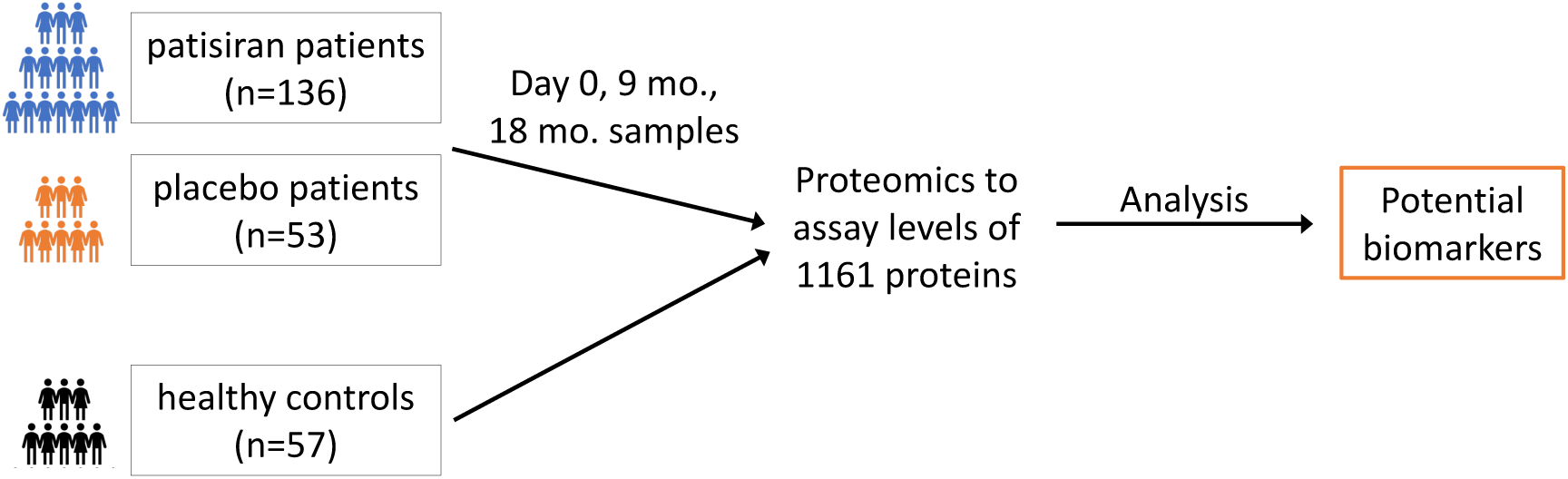
Overview of study design and samples analysed. mo.=months.

### Multiple plasma proteins differ significantly between placebo- and patisiran-treated patients

Plasma levels of 1161 unique proteins were analysed using a proximity extension assay. A linear mixed model was used to determine the impact of patisiran treatment on the time profile of each protein’s plasma level by analysing levels at baseline and 9 and 18 months. A total of 66 proteins were found to show a significant change in levels in placebo *vs* patisiran-treated patients over time (p<4·18×10^−5^; applying Bonferroni correction), of which NfL was the most significant (figure 2; p=3·95×10^−21^). This analysis revealed that upon treatment with patisiran levels of some proteins increased (eg, neutral ceramidase) whereas levels of other proteins decreased (eg, NfL, R-spondin 3, coiled-coil domain containing 80). Many of the proteins affected by patisiran treatment have not been described as biomarkers (table 1; appendix); however, one previously described biomarker of cardiac health, *N*-terminal pro-brain natriuretic peptide (NT-proBNP) showed a significant decrease (p=7·02×10^−14^) upon patisiran treatment. The NT-proBNP levels predicted by the proximity extension assay data and linear mixed model were consistent with those measured using an orthogonal method in the APOLLO study.^6^

**Figure 2:**
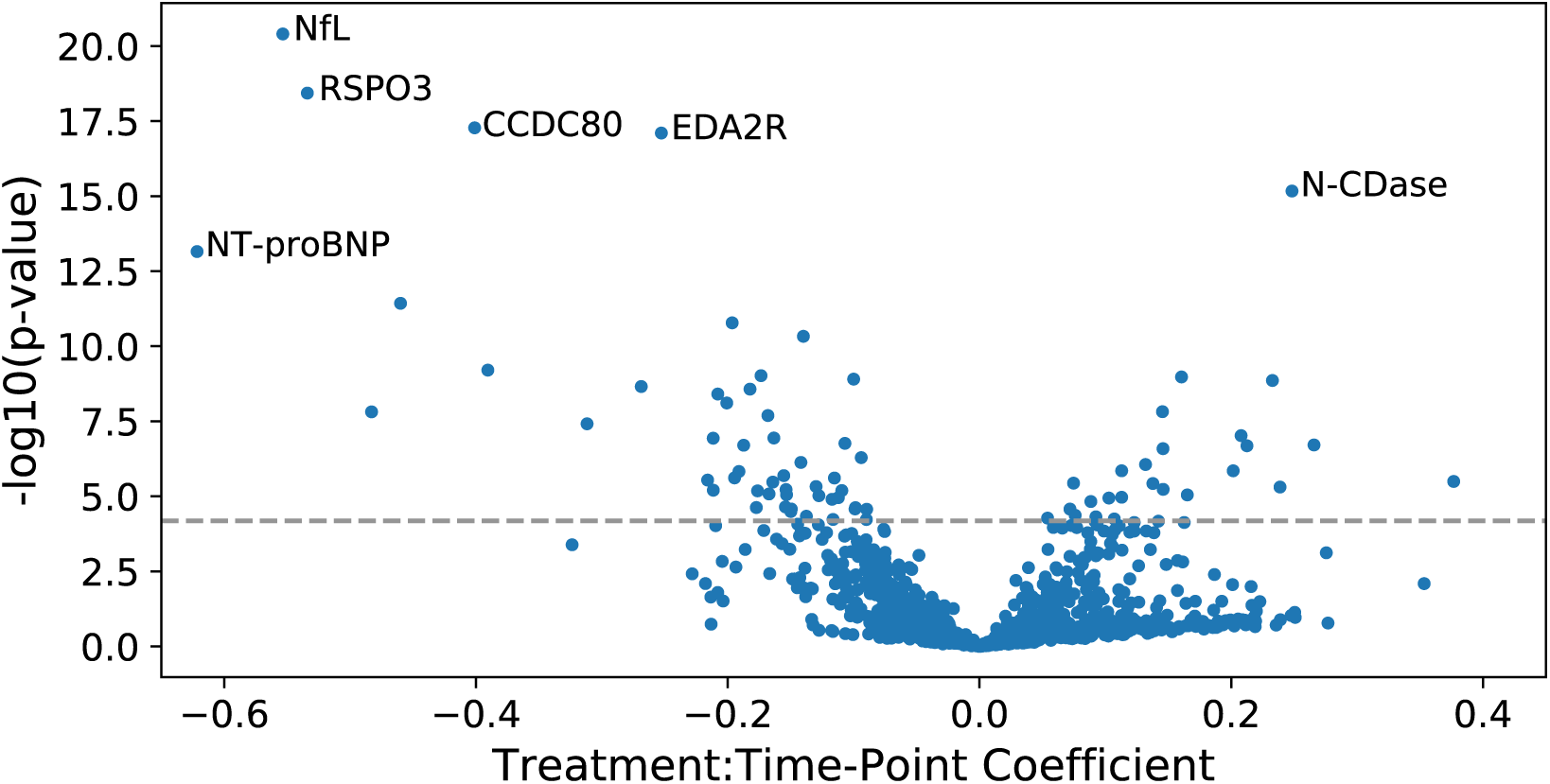
Proteins identified that have a change corresponding to patisiran treatment (relative to placebo) over 18 months. Proteins are shown here as a volcano plot, with the strength of the association on the y-axis (–log_10_[p value]) and the effect size on the x-axis (shown as the treatment×time coefficient from the model). A full list of the 66 most enriched proteins can be found in table 1; appendix. CCDC80=coiled-coil domain containing 80; EDA2R=ectodysplasin A2 receptor; N-CDase=neutral ceramidase; NfL=neurofilament light chain; NT-proBNP=*N*-terminal prohormone of brain-type natriuretic peptide; RSPO3=R-spondin 3.

### Effect of disease progression and treatment on overall plasma proteome signature

Principal component analysis was used to elucidate whether there was a systemic plasma proteomic shift in patients with hATTR amyloidosis, and whether there was a subsequent reversal following treatment with patisiran. The 66 most significant proteins found in the mixed model analysis (table 1; appendix) were compared in samples from healthy controls and baseline samples from patients with hATTR amyloidosis. Projecting each individual’s data onto the first two principal components that account for the most variance revealed a distinct separation between healthy controls and patients with hATTR amyloidosis with polyneuropathy at the APOLLO baseline (figure 3A), with most of the separation being driven by the first principal component (PC1). The proteomes of placebo-treated patients at 18 months were projected onto the same principal components resulting in a leftward movement along PC1 of the 99·9% confidence ellipse, suggesting that the proteome of placebo-treated patients was further separating from healthy controls (figure 3B). At 18 months, it was found that plasma proteomes of patisiran-treated patients were more similar to those of healthy subjects compared with placebo-treated patients (figure 3C). Since there is significant heterogeneity in the proteome of patients at baseline, individual patient trajectories were also captured in 66-dimensional space. For each patient, the course of their disease progression or reversal was defined using two metrics, one representing the rate (ϕ) and a second representing whether the proteome moved away from or towards the healthy controls (θ; see figure 3D legend). Plotting these two parameters clearly separated patisiran- and placebo-treated patients (figure 3E), emphasising the distinct differences in the proteomes of the two groups at 18 months.

**Figure 3:**
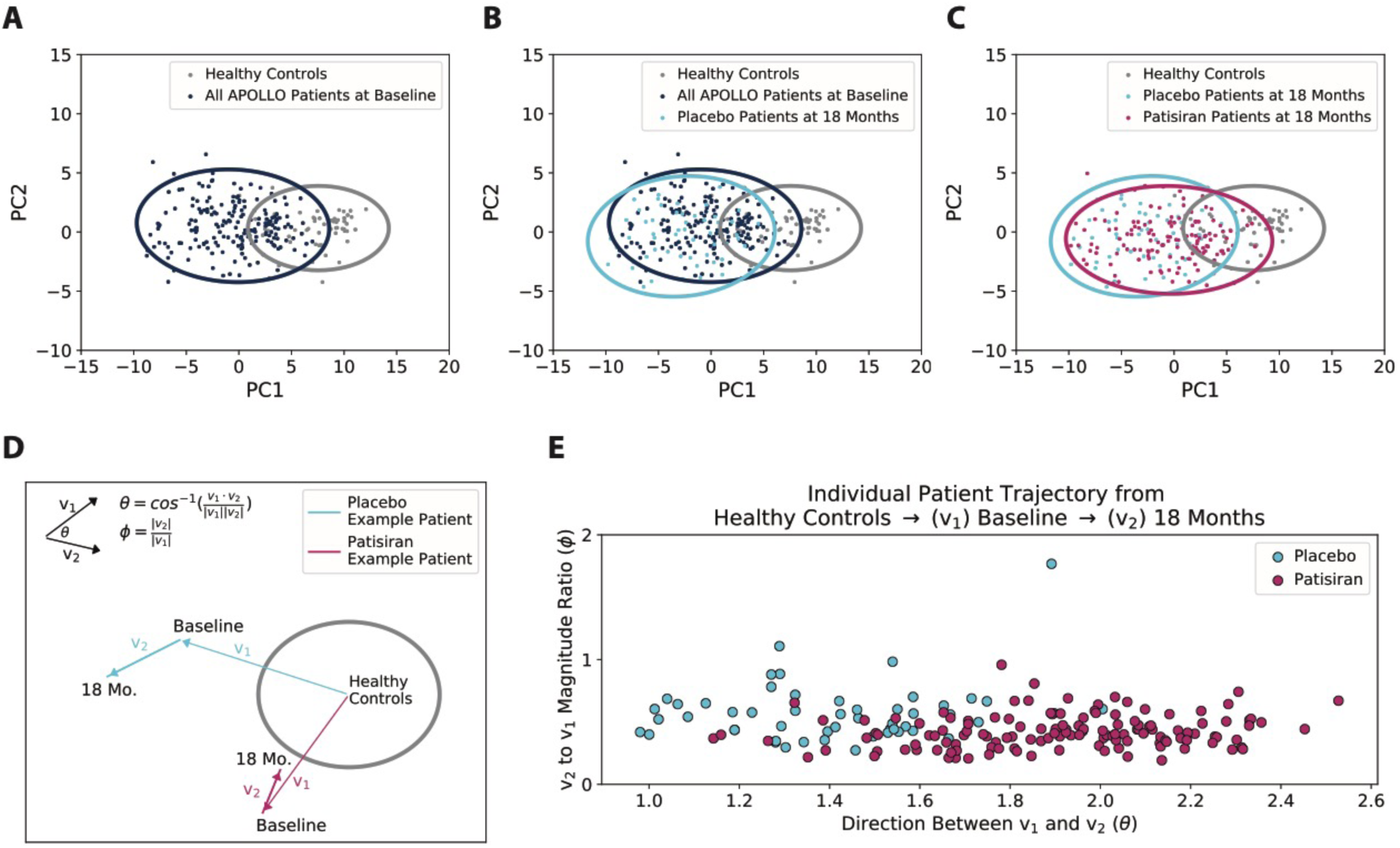
Global changes in plasma proteomes observed with the proteome of patisiran-treated patients trending towards that of healthy controls at 18 months. (A) A subset of the measured proteins was used to project the differences between patients with hATTR amyloidosis and healthy controls at baseline onto two principal components (PC1 and PC2) that most explained the difference in the data sets; analysis of (B) placebo-treated patients at 18 months and (C) patisiran-treated patients at 18 months is shown in the same PC1 and PC2 space. (D) Illustrative diagram depicting v1 and v2 as well as ϕ and θ. For each patient, a vector (v1) from the mean healthy patient to baseline and a second vector (v2) from baseline to month 18 were used to compute two metrics, one for the ratio of the magnitude of v2 compared to v1 denoted as ϕ, and θ as the angle between the two vectors. Here ϕ measures the rate of disease progression or reversal and θ measures the directionality of the proteome (defined by whether the proteome moves away or towards the healthy controls). (E) Individual patient trajectories are shown, separated by whether patients were on placebo or patisiran treatment. hATTR=hereditary transthyretin-mediated; Mo.=months; PC=principal component.

### Plasma NfL is increased in patients with hATTR amyloidosis with polyneuropathy and decreases with patisiran treatment

NfL was the most significantly changed protein in the analysis when comparing placebo- and patisiran-treated patients with hATTR amyloidosis with polyneuropathy and was therefore investigated in more detail. Patients diagnosed with hATTR amyloidosis with polyneuropathy had greater than four-fold higher levels of NfL in their plasma at baseline relative to healthy controls (figure 4A; log_2_ scale). As expected, plasma NfL levels at baseline did not differ between the patisiran and placebo groups. The patisiran-treated group showed a significant decline in plasma NfL levels at 9 months that was sustained at 18 months whereas plasma NfL levels in the placebo group increased at 9 months relative to baseline and this level was sustained at 18 months (figure 4B). At 18 months, placebo-treated patients had two-fold higher NfL plasma levels than patients treated with patisiran. Treatment with patisiran significantly lowered NfL levels in patients with hATTR amyloidosis towards levels observed in healthy controls.

**Figure 4:**
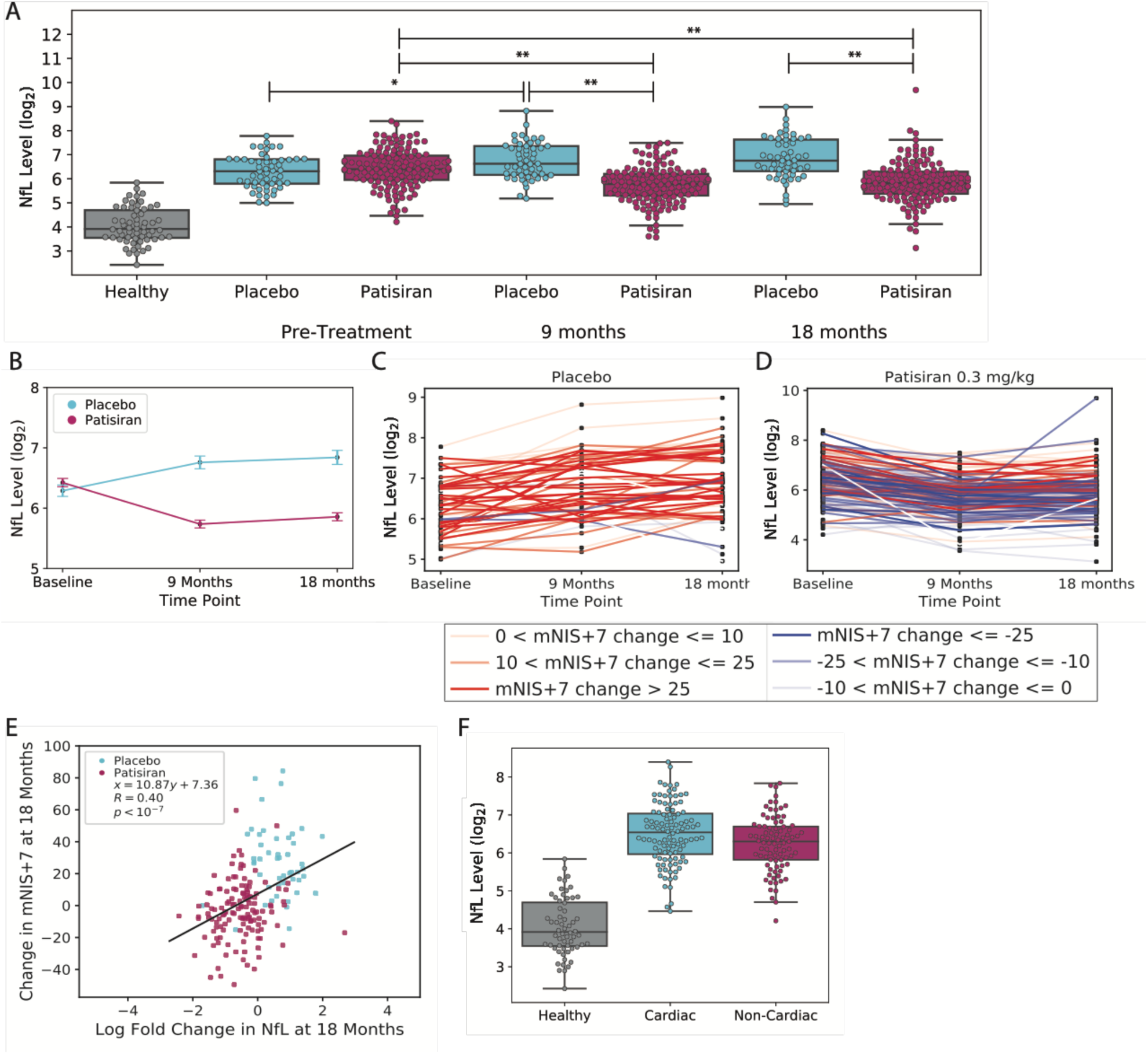
Change over time in NfL in patients treated with placebo and patisiran. (A) Levels of NfL (NPX values which are on a log_2_ scale) in healthy controls and placebo- or patisiran-treated patients at baseline, 9 months, or 18 months. (B) Mean±standard error of NfL levels in patisiran- and placebo-treated patients at baseline and 9 and 18 months. Trajectories of individual patients on (C) placebo or (D) patisiran over time, colour-coded by their corresponding worsening (change in mNIS+7>0; red) or improvement (change in mNIS+7<0; blue) in mNIS+7 from baseline to 18 months (E) Correlation between change in NfL levels from baseline to 18 months and the corresponding change in mNIS+7 coloured by treatment. (F) Levels of NfL separated by whether patients were in the predefined cardiac subset of APOLLO or not, at baseline. *p<0·01, **p<0·001. NfL=neurofilament light chain; NPX= normalised protein expression; mNIS+7=modified Neuropathy Impairment Score+7.

To assess variability between individuals, plasma NfL levels were plotted over time for each patient receiving placebo (figure 4C) or patisiran (figure 4D) treatment. Consistent with the general trend in the previous analysis, patients receiving placebo showed increasing plasma NfL levels over time, whereas patients receiving patisiran demonstrated decreasing plasma NfL levels. Changes in plasma NfL levels were correlated with mNIS+7 score at 18 months (*R*=0·4; figure 4E) indicating that decreasing plasma NfL levels are associated with an improvement in polyneuropathy. Notably, no correlation was observed between plasma NfL levels and mNIS+7 score at baseline (figure 1; appendix). Additionally, patients belonging to the pre-specified cardiac subpopulation of the APOLLO study (defined as baseline left ventricular wall thickness ≥13 mm in the absence of a history of aortic valve disease or hypertension)^6^ saw similar elevation of plasma NfL levels (figure 4F) to patients who only had symptoms of polyneuropathy, suggesting that NfL can serve as a biomarker of polyneuropathy in patients regardless of cardiac involvement.

The initial NfL measurements were validated using a widely used ultrasensitive single molecule quantitative assay to provide absolute plasma concentrations of NfL in healthy controls and hATTR amyloidosis patient samples from APOLLO at baseline and 18 months (figure 5). There was a very strong correlation between the two assays used to measure plasma NfL levels (figure 5A; *R*=0·96). Using the quantitative assay, the mean NfL level in healthy individuals was 16·3 pg/mL, consistent with published reports (SD 12·0 pg/mL; figure 5B).^17^ Patients with hATTR amyloidosis with polyneuropathy had plasma NfL levels that were approximately four-fold higher at baseline, 69·4 pg/mL (SD 42·1 pg/mL), which, after 18 months, decreased following patisiran treatment (48·8 pg/mL ± 29·9 pg/mL) and increased in the placebo group (99·5 pg/mL ± 60·1 pg/mL). One outlier with NfL plasma levels of 747 pg/mL in the patisiran-treated group was excluded from the calculations (figure 5B and 5E) as this individual experienced a study-unrelated cerebral infarct at month 17 that may have caused the observed elevations of NfL. Consistent with the initial plasma NfL measurements (figure 4), measurements using the quantitative assay demonstrated a strong correlation between change in mNIS+7 and change in plasma NfL levels at 18 months for each patient (*R*=0·43; figure 5C). A receiver operator curve determined that plasma NfL levels could discriminate between healthy individuals and patients with hATTR amyloidosis with polyneuropathy (area under the curve of 0·956; figure 5D) and the distribution of each was plotted (figure 5E). Based on the current data, 37 pg/mL can be considered a conservative cut-off to distinguish between healthy patients and patients with hATTR amyloidosis, at which the false positive rate is 3·6% and the true positive rate is 84·9%.

**Figure 5:**
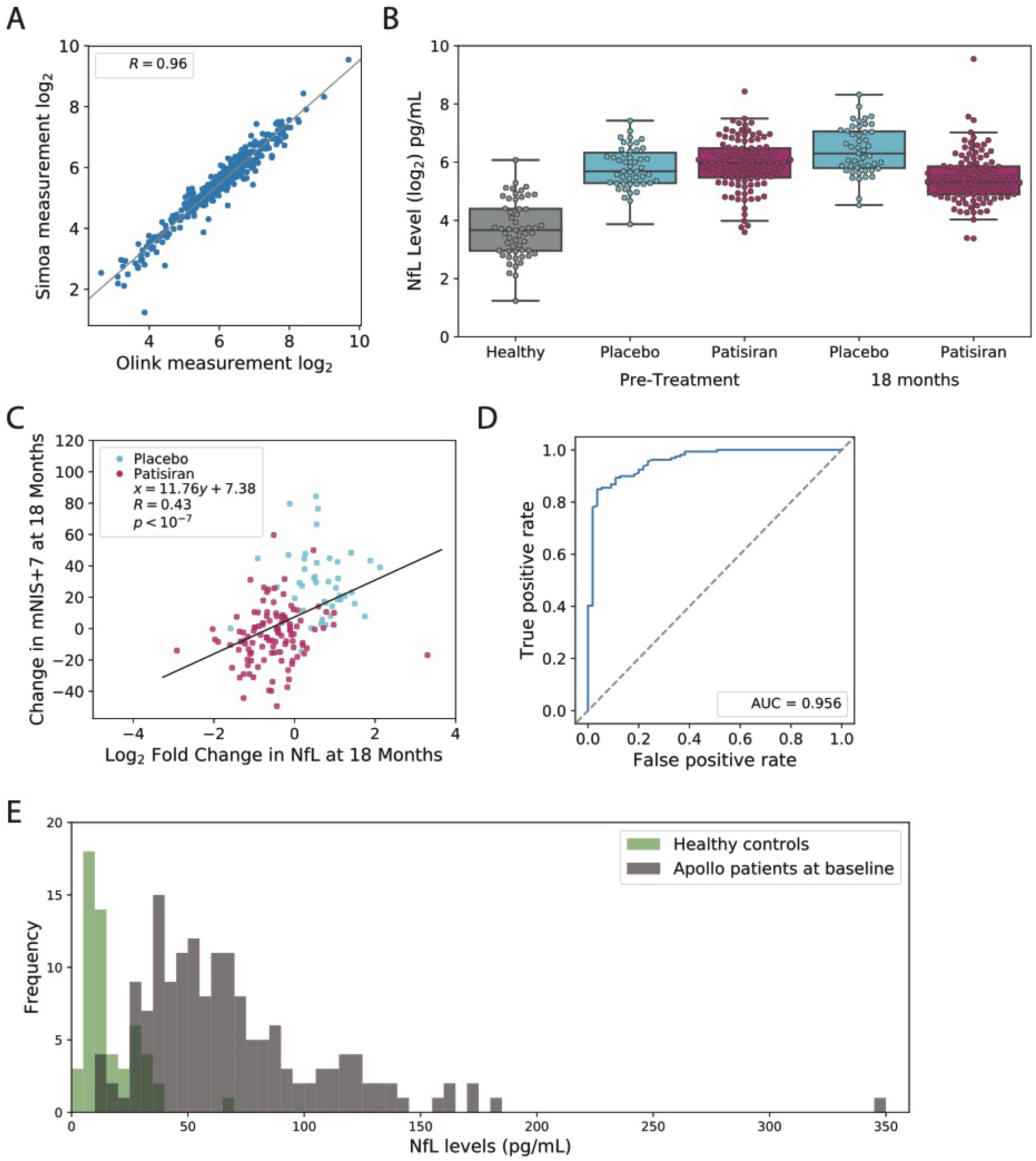
Quantitative measurement of NfL confirm previous findings and show potential of plasma NfL levels to distinguish between healthy patients and patients with hATTR amyloidosis with polyneuropathy. (A) Correlation between Olink^®^ and Quanterix Simoa^®^ platform is 0·96. (B) Levels of NfL in healthy controls and placebo- or patisiran-treated patients at baseline or 18 months. (C) Correlation between change in NfL levels from baseline to 18 months and the corresponding change in mNIS+7 coloured by treatment. (D) Receiver operator characteristic curve analysis of NfL plasma levels in healthy patients *vs* patients with hATTR amyloidosis with polyneuropathy. The area under the curve is 0·956. (E) Histograms showing the distributions of NfL concentrations in healthy controls (green) and patients with hATTR amyloidosis at baseline (dark grey). hATTR=hereditary transthyretin-mediated; NfL=neurofilament light chain; mNIS+7=modified Neuropathy Impairment Score+7.

## Discussion

Plasma proteomics and the identification of minimally invasive biomarkers are emerging as integral parts of modern drug discovery and clinical development. In an effort to leverage this approach, the plasma proteomes of patients with hATTR amyloidosis with polyneuropathy were investigated over time in the largest clinical proteomic study of this disease to date. Proteomic analyses of patient plasma samples demonstrated that patisiran treatment resulted in a general shift of patients’ proteomes towards those of healthy individuals compared with patients who received placebo treatment. This finding suggests that the plasma protein milieu is reflective of hATTR amyloidosis disease progression and response to treatment. Across >1000 unique proteins that were assessed from plasma samples collected in the APOLLO study, 66 proteins were found to exhibit a significantly different plasma level time profile in patisiran-compared with placebo-treated patients over the course of 18 months. Of these, NfL was identified as the protein most significantly different between the two groups. Collectively these findings demonstrate that patient proteome changes are detectable following treatment, as well as support the identification of proteins that have both the potential to serve as biomarkers and offer insights into hATTR amyloidosis disease biology.

Of highest clinical interest are the high plasma NfL levels detected in patients with hATTR amyloidosis with polyneuropathy, and the subsequent decrease of these following patisiran treatment. This is important given the emerging role of NfL as a general biomarker of central and peripheral nervous system dysfunction. NfL is an integral component of the axonal structure of neurons and has been described extensively as a biomarker of neuroaxonal injury across many central nervous system diseases including multiple sclerosis,^18^ Alzheimer’s,^19^ and Huntington’s,^16^ as well as peripheral nervous system diseases such as vasculitis,^20^ chronic inflammatory demyelinating neuropathy (CIDP),^21^ Guillain–Barré syndrome,^22^ and Charcot–Marie–Tooth disease (CMT).^23^ Mutations in NfL itself can also lead to CMT disease, a group of inherited peripheral neuropathies, indicating that NfL plays a crucial role in peripheral nerve function.^24^ Elevated NfL levels in these diseases are thought to be caused by the release of NfL into circulation from damaged neurons^25^ and it has been identified in both cerebrospinal fluid^26^ and blood.^27^ In this study the mean NfL levels described in patients with hATTR amyloidosis with polyneuropathy (69·4 pg/mL) were higher than those observed for other peripheral nerve disorders (CIDP 42 pg/mL, CMT 26 pg/mL)^21,23^ and consistent with a previous report in patients with hATTR amyloidosis (58·1 pg/mL).^28^

Patisiran treatment lowers *TTR* messenger RNA levels (and therefore circulating TTR protein), potentially resulting in a reduction of amyloid burden, which is hypothesised to halt or reduce neuronal damage.^15^ This hypothesis was supported by the improvement of polyneuropathy seen in patients treated with patisiran in the APOLLO study. The current study suggests that a patisiran-mediated reduction in nerve damage also causes a reduction in plasma levels of NfL. Reductions in NfL levels observed following only 9 months of patisiran treatment indicate that treatment reduces hATTR amyloidosis-associated neuronal damage within a short timeframe. While the reduction in NfL levels was sustained for a further 9 months of patisiran treatment, the lack of a further meaningful reduction between months 9 and 18 towards levels detected in healthy individuals is not in alignment with the clinical results from APOLLO, in which there was an additional improvement in mNIS+7 between month 9 and 18. Further investigation would be necessary to explain this discrepancy.

The correlation observed between change in mNIS+7 score and change in plasma NfL levels over 18 months indicates that decreasing levels of NfL detected in patients treated with patisiran are also associated with improvement in polyneuropathy. There was, however, no correlation between NfL levels and mNIS+7 score at baseline. One explanation for this observation is that significant heterogeneity exists in inter-patient plasma NfL levels, while intra-patient NfL changes over time may be more informative regarding changes in disease status.

Notably, levels of NfL were elevated at baseline in patients with and without evidence of cardiac amyloid involvement, suggesting that NfL may be useful as a biomarker of polyneuropathy in patients previously diagnosed with hATTR amyloidosis with cardiomyopathy. An investigation of plasma NfL levels in patients with predominantly cardiomyopathic phenotype may further support hATTR amyloidosis as a single multisystem disease, regardless of *TTR* mutation or initial presenting symptoms.

In addition to monitoring disease progression and treatment response, NfL may serve as a potential diagnostic biomarker or as an indicator for the transition from an asymptomatic carrier to a symptomatic patient. Studies reported in the literature suggest NfL can serve as a prognostic biomarker in pre-symptomatic amyotrophic lateral sclerosis^29^ and Alzheimer’s disease.^30^ The data described here suggest that a plasma NfL concentration of 37 pg/mL may represent a clinically meaningful threshold to aid the diagnosis of neuronal damage in hATTR amyloidosis with polyneuropathy, supplementing clinical assessments that are already being used. Due to the heterogeneity observed in patients with hATTR amyloidosis with polyneuropathy, the amount of change in NfL levels over time may prove to be a more sensitive indicator of neuronal damage than absolute levels. In plasma samples collected from the phase 3 APOLLO study, an average increase of 30 pg/mL was observed in the placebo group over the course of 18 months, indicating that NfL may also serve as a biomarker of disease progression.

In conclusion, this study represents the first system-wide proteomics interrogation of response to an RNAi therapeutic in humans. The analysis of samples collected from APOLLO patients has led to an improved understanding of polyneuropathy progression in patients with hATTR amyloidosis. NfL has been identified as a protein that is differentially expressed in patients with hATTR amyloidosis *vs* healthy individuals. Changes in plasma NfL levels are also seen in response to patisiran treatment over time, highlighting the potential of this protein to serve as a biomarker in several capacities, including as an early diagnostic biomarker of hATTR amyloidosis or a predictor of when disease may become manifest in asymptomatic carriers of pathogenic *TTR* mutations, and as an objective marker for monitoring disease progression and/or reversal over time. The results from this comprehensive, proteomic analysis using samples from a placebo-controlled study have (1) identified NfL as a potential multi-purpose biomarker for hATTR amyloidosis, (2) provided new insights into hATTR amyloidosis disease biology, and (3) enabled the first system-wide proteomics interrogation of response to an RNAi therapeutic in humans.

## Data Availability

Not applicable

## Research in context

### Evidence before this study

A PubMed search was carried out for articles published up to 25 October 2019 on circulating biomarkers in transthyretin amyloidosis using the terms “(([TTR] OR [transthyretin]) AND ([amyloidosis] OR [polyneuropathy]) AND [biomarkers] AND ([blood] OR [serum] OR [plasma]))”. Only two studies from the same research group were identified from this search. These studies analysed the concentrations of plasma proteins in healthy controls, patients with polyneuropathy, or patients with cardiomyopathy caused by hereditary transthyretin-mediated (hATTR) amyloidosis or wild-type transthyretin-mediated amyloidosis, using multiple-reaction monitoring mass spectrometry. Study cohorts were small, with only 8–10 individuals included in each study and a total of 160 different proteins measured. Additionally, these studies did not examine changes in plasma levels of each protein over time, or in response to treatment.

A PubMed search was also completed for articles published up to 25 October 2019 on NfL in ATTR amyloidosis by use of the terms “(([TTR] OR [transthyretin]) AND ([amyloidosis] OR [polyneuropathy]) AND ([neurofilament light chain] or [NfL] or [NEFL]))”. Only one previous study was identified which examined plasma NfL levels in patients with hATTR amyloidosis. This study found elevated levels of NfL in 73 patients with hATTR amyloidosis; however, it did not analyse change in plasma NfL levels over time or after initiation of a therapeutic and it did not investigate any other circulating proteins.

### Added value of this study

To our knowledge, this study is the largest proteomic study to date in patients with hATTR amyloidosis with polyneuropathy, analysing plasma levels of >1000 proteins in samples from 246 individuals. Furthermore, this is the first study to analyse changes in the plasma proteome occurring in response to an RNAi therapeutic in humans. These findings provide additional evidence to suggest that NfL may be an important biomarker for patients with hATTR amyloidosis with polyneuropathy. Additionally, they provide new evidence that demonstrates the potential of NfL as a biomarker to monitor disease progression/reversal and response to treatment in patients with hATTR amyloidosis with polyneuropathy.

### Implications of all the available evidence

Measurement of plasma NfL levels may serve as a non-invasive, complementary approach to facilitate earlier diagnosis, and monitor disease progression and treatment response in patients with hATTR amyloidosis with polyneuropathy. However, further evidence will be important to confirm these findings. Additional proteins were identified that may also serve as biomarkers in hATTR amyloidosis with polyneuropathy, although their involvement in disease biology will require further investigation.

### Contributors

ST(Ticau), GVS, AC, KF, AV, and PN designed the study. Data was analysed by ST(Ticau), GVS, and ST(Tsour) and interpreted by all authors. ST(Ticau), GVS and ST(Tsour) created the figures. All authors were involved in the writing of the report.

## Acknowledgements

The authors would like to thank the patients and their families involved in the APOLLO study for their valued contribution to this study. This study was funded by Alnylam Pharmaceuticals Inc., USA. Editorial support was provided by Ed Childs, PhD, of Adelphi Communications Ltd, Macclesfield, UK, in accordance with the Good Publication Practice (GPP3) guidelines, funded by Alnylam Pharmaceuticals Inc, USA.

## Availability of data and materials

The datasets generated and analysed during the current study are not publicly available.

---

**Table S1:** Comprehensive list of all proteins that significantly changed (Bonferroni threshold p<4·18×10^−5^) found using a linear mixed model to compare placebo- and patisiran-treated patients over time·beta coefficient of time×treatment term and –log_10_(p value) are listed

| Protein | Beta coefficient | $-\log_{10}$ (p value) |
| --- | --- | --- |
| NfL | 0.55 | 20.40 |
| RSPO3 | 0.53 | 18.43 |
| CCDC80 | 0.40 | 17.28 |
| EDA2R | 0.25 | 17.10 |
| N-CDase | -0.25 | 15.17 |
| NT-proBNP | 0.62 | 13.15 |
|  | 0.46 | 11.43 |
| WNT9A | 0.20 | 10.78 |
| SMOC2 | 0.14 | 10.33 |
| PTN | 0.39 | 9.20 |
| HGF | 0.17 | 9.02 |
|  | 0.14 | 6.13 |
| NELL1 | -0.16 | 8.98 |
| DCN | 0.10 | 8.90 |
| ARSA | -0.23 | 8.86 |
| KLK4 | 0.27 | 8.66 |
| GPC1 | 0·18 | 8·57 |
| SFRP | 0·21 | 8·41 |
| DRAXIN | 0·20 | 8·11 |
| IL-18R1 | −0·15 | 7·82 |
| BNP | 0·48 | 7·81 |
| GFR-alpha1 | 0·17 | 7·69 |
| CXCL9 | 0·31 | 7·42 |
| TIMD4 | −0·21 | 7·02 |
| RSPO1 | 0·16 | 6·94 |
| MYOC | 0·21 | 6·94 |
| WFDC2 | 0·11 | 6·76 |
| GUSB | −0·27 | 6·71 |
| HSPB6 | 0·19 | 6·70 |
| MFGE8 | −0·21 | 6·68 |
| SMPD1 | −0·15 | 6·59 |
| FLT1 | 0·09 | 6·29 |
| CTSD | −0·13 | 6·06 |
| CD209 | −0·11 | 5·85 |
| LDLR | −0·20 | 5·85 |
| NOV | 0·19 | 5·83 |
| CD160 | 0·16 | 5·69 |
| PI3 | 0·19 | 5·61 |
| SMOC1 | 0·12 | 5·61 |
| TFPI-2 | 0·22 | 5·54 |
| LEP | −0·38 | 5·49 |
| TNFRSF19 | 0·16 | 5·47 |
| ERBB3 | −0·07 | 5·44 |
| DUA | −0·14 | 5·43 |
| IL-4RA | 0·13 | 5·33 |
| REN | −0·24 | 5·31 |
| CES2 | −0·15 | 5·23 |
| CR2 | 0·15 | 5·23 |
| PSG1 | 0·21 | 5·20 |
| FGF-BP1 | 0·11 | 5·19 |
| OPN | 0·18 | 5·18 |
| TFF3 | 0·17 | 5·08 |
| ANGPT2 | 0·15 | 5·05 |
| CCL24 | −0·17 | 5·05 |
| LAYN | 0·13 | 5·02 |
| FUCA1 | −0·11 | 4·97 |
| AXL | 0·11 | 4·95 |
| TLR3 | −0·10 | 4·94 |
| SCARB2 | 0·12 | 4·90 |
| B4GAT1 | −0·09 | 4·83 |
| SORCS2 | 0·15 | 4·65 |
| CD300E | 0·18 | 4·63 |
| CD27 | 0·10 | 4·62 |
| SPON1 | 0·10 | 4·58 |
| IGFBP-2 | 0.15 | 4.58 |
| DNER | -0.07 | 4.57 |
| RARRES1 | 0.09 | 4.57 |
| Dkk-4 | 0.15 | 4.49 |
| **Table S1: Comprehensive list of all proteins that significantly changed (Bonferroni threshold $p < 4.18 \times 10^{-5}$) found using a linear mixed model to compare placebo- and patisiran-treated patients over time. beta coefficient of time × treatment term and $-\log_{10}(p \text{ value})$ are listed** |  |  |

**Figure S1:**
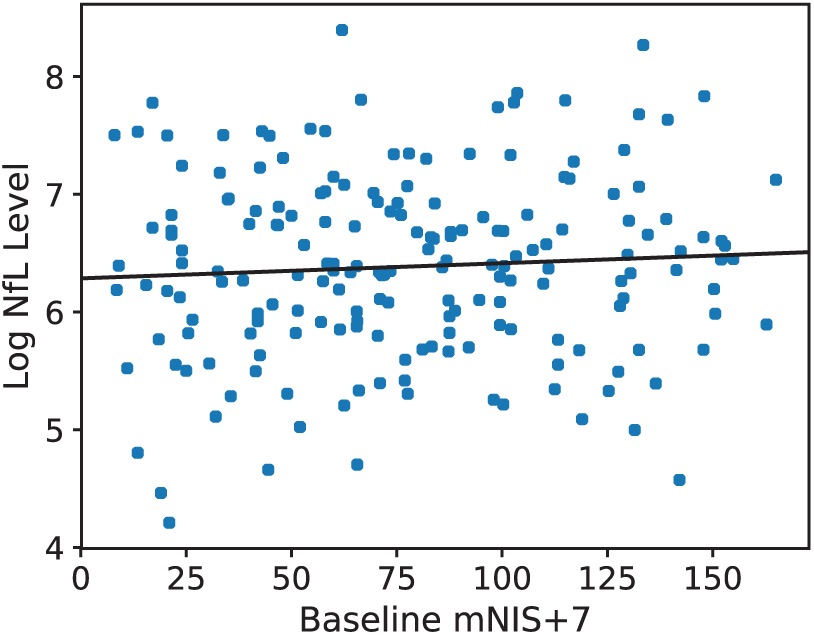
Association between NfL levels and mNIS+7 at baseline. NfL=neurofilament light chain; mNIS+7=modified Neuropathy Impairment Score+7.

